# Self-harm during the early period of the COVID-19 Pandemic in England: comparative trend analysis of hospital presentations

**DOI:** 10.1101/2020.11.25.20238030

**Authors:** Keith Hawton, Deborah Casey, Elizabeth Bale, Fiona Brand, Jennifer Ness, Keith Waters, Sam Kelly, Galit Geulayov

## Abstract

**Background:** The COVID-19 pandemic and public health measures necessary to address it may have major effects on mental health, including on self-harm. We have used well-established monitoring systems in two hospitals in England to investigate trends in self-harm presentations to hospitals during the early period of the pandemic.

**Method:** Data collected in Oxford and Derby on patients aged 18 years and over who received a psychosocial assessment after presenting to the emergency departments following self-harm were used to compare trends during the three-month period following lockdown in the UK (23^rd^ March 2020) to the period preceding lockdown and the equivalent period in 2019.

**Results:** During the 12 weeks following introduction of lockdown restrictions there was a large reduction in the number of self-harm presentations to hospitals by individuals aged 18 years and over compared to the pre-lockdown weeks in 2020 (mean weekly reduction of 13.5 (95% CI 5.6 - 21.4) and the equivalent period in 2019 (mean weekly reduction of 18.0 (95% CI 13.9 - 22.1). The reduction was greater in females than males, occurred in all age groups, with a larger reduction in presentations following self-poisoning than self-injury.

**Conclusions:** A substantial decline in hospital presentations for self-harm occurred during the three months following the introduction of lockdown restrictions. Reasons could include a reduction in self-harm at the community level and individuals avoiding presenting to hospital following self-harm. Longer-term monitoring of self-harm behaviour during the pandemic is essential, together with efforts to encourage help-seeking and the modification of care provision.

## INTRODUCTION

The challenges to health and society posed by the COVID-19 pandemic are huge. Understandably, while the initial focus was on physical health and prevention of spread of the disease and of deaths, attention has also increasingly turned to the potential mental health consequences of the pandemic (Holmes et al., 2020), including the possible impacts on suicidal behavior (Gunnell et al., 2020; Reger et al., 2020) (Niederkrotenthaler et al., 2020). Concerns have been expressed about the psychological consequences of the necessary public health measures, including lockdown and social distancing (Brooks et al., 2020; Pierce et al., 2020). Lockdown was introduced in the UK on March 23^rd^ 2020, easing of lockdown in England being announced on May11^th^ 2020. Attention has also turned to what the potential impacts of the longer-term consequences of the pandemic may be for mental health, including, for example, those resulting from unemployment, financial problems, reduced access to schooling, and bereavement (Gunnell et al., 2020; Holmes et al., 2020; Niederkrotenthaler et al., 2020).

Suicide and self-harm are tangible measures of mental health problems. Both are known to be affected by social and economic factors (Hawton et al., 2014; Turecki et al., 2019). In this study we have used well-established monitoring systems to investigate trends in self-harm presentations to hospitals in England, particularly focusing on the three-month period following lockdown in the UK (from March 23^rd^, 2020), and including comparison with the pattern preceding lockdown in 2020 and the equivalent period in 2019. We have examined trends by gender, age and method of self-harm.

## METHOD

### Patients

We used information on all patients aged 18 years and over presenting to emergency departments following self-harm who received a psychosocial assessment (of their problems, needs and risks). Self-harm is defined as intentional non-fatal acts of self-poisoning or self-injury, irrespective of the type of motivation, including degree of suicidal intent (Hawton, 2003). We used data from two centres participating in the Multicentre Study of Self-harm in England (Oxford and Derby) for the periods 6^th^ January to 14^th^ June 2020. This period included the seven weeks following lockdown that started on Monday 23^rd^ March 2020 and the first five weeks following the first easing of lockdown on May 11th. We have also included data for the equivalent period in 2019 to assess for possible period effects.

The information was based on assessed presentations rather than all presentations as there was limited access to information on non-assessed individuals who attended the ED in one of the study centres due to lockdown restrictions. Our data from previous years show that assessment rate in these two centres is around 75% of presentations following self-harm. We have no reason to believe that this has changed during the study period.

### Data collection

In Oxford, data were collected from monitoring forms completed by clinical staff after assessing patients who presented with self-harm. In Derby, data were extracted from electronic patient records completed by the liaison psychiatry team following assessment of patients who had presented following self-harm.

The datasets consisted of de-identified variables, in order to maintain patient confidentiality, consisting of researcher-generated episode numbers, date of presentation to the emergency department, broad age groups (18-34 years, 35-54 years and 55 years and over), and general method of self-harm (self-poisoning, self-injury, and both self-poisoning and self-injury in the same episode).

### Statistical analyses

The data are presented in terms of numbers of presentations by time period, age group, gender, and method of self-harm. We have examined the difference in mean assessed presentations per calendar week following introduction of lockdown restrictions compared with the preceding weeks in 2020 and also compared these respective periods with the equivalent periods in 2019. We present the differences as means with 95% confidence intervals for each of these comparisons. These differences were tested using independent samples t-tests, not assuming equal variance. We also used regression analysis to examine differences in incidence of self-harm after lockdown compared with beforehand by weekly numbers. To accommodate for over-dispersion of data, as indicated by a goodness of fit test, we used Negative Binomial regression. The findings of these analyses are presented as incidence rate ratios (IRRs).

We have compared changes in numbers of presentations between males and females, including calculating the gender ratio before and after lockdown. We present IRRs with 95% confidence intervals, using negative binomial regression.

To assess the effect of lockdown by age group, we have examined the changes in presentations to hospital in three broad age groups (18-34 years, 35-54 year and 55 years and over) and have presented IRRs from the negative binomial regression analysis.

We have examined the effect of lockdown by broad method of self-harm, presenting IRRs from a negative binomial regression model.

Analyses were conducted using SPSS v25 and Stata v14.2.

### Ethical approval

The monitoring systems in Oxford and Derby have Health Research Authority and National Health Service (NHS) Research Ethics Committee approval. The two monitoring systems are fully compliant with the Data Protection Act (1998) and have approval under Section 251 of the NHS Act (2006) to collect patient-identifiable information without patient consent.

## RESULTS

### Sample

During the study period 6^th^ January 2020 to 14^th^ June 2020 and the equivalent period in 2019 there were 1957 episodes of self-harm by individuals aged 18 years and over who received psychosocial assessments (985 presentations in Oxford and 972 presentations in Derby). These are henceforth referred to as ‘presentations’. These included 1186 (60.6%) by females and 771 (39.4%) by males

### Changes in presentation associated with lockdown

During the period 6^th^ January to 14^th^ June 2020 there were 854 hospital presentations (431 in Oxford and 423 in Derby) compared with 1103 during the same period in 2019 (554 in Oxford and 549 in Derby).

The weekly numbers of presentations during the period following the beginning of lockdown (March 23^rd^ 2020 to 14^th^ June 2020) were markedly lower than the weekly numbers observed during the preceding period in 2020 and also compared to the equivalent period in 2019. Numbers rose somewhat during the weeks following easing of lockdown in England on12th May, but still remained lower than during the period prior to lockdown and the equivalent period in 2019. (Figure 1). Of note is the observation that the reduction of self-harm presentations may have begun in the week before lockdown (The Secretary of State for Health announced the forthcoming lockdown on March 16^th^).

**Figure 1.**
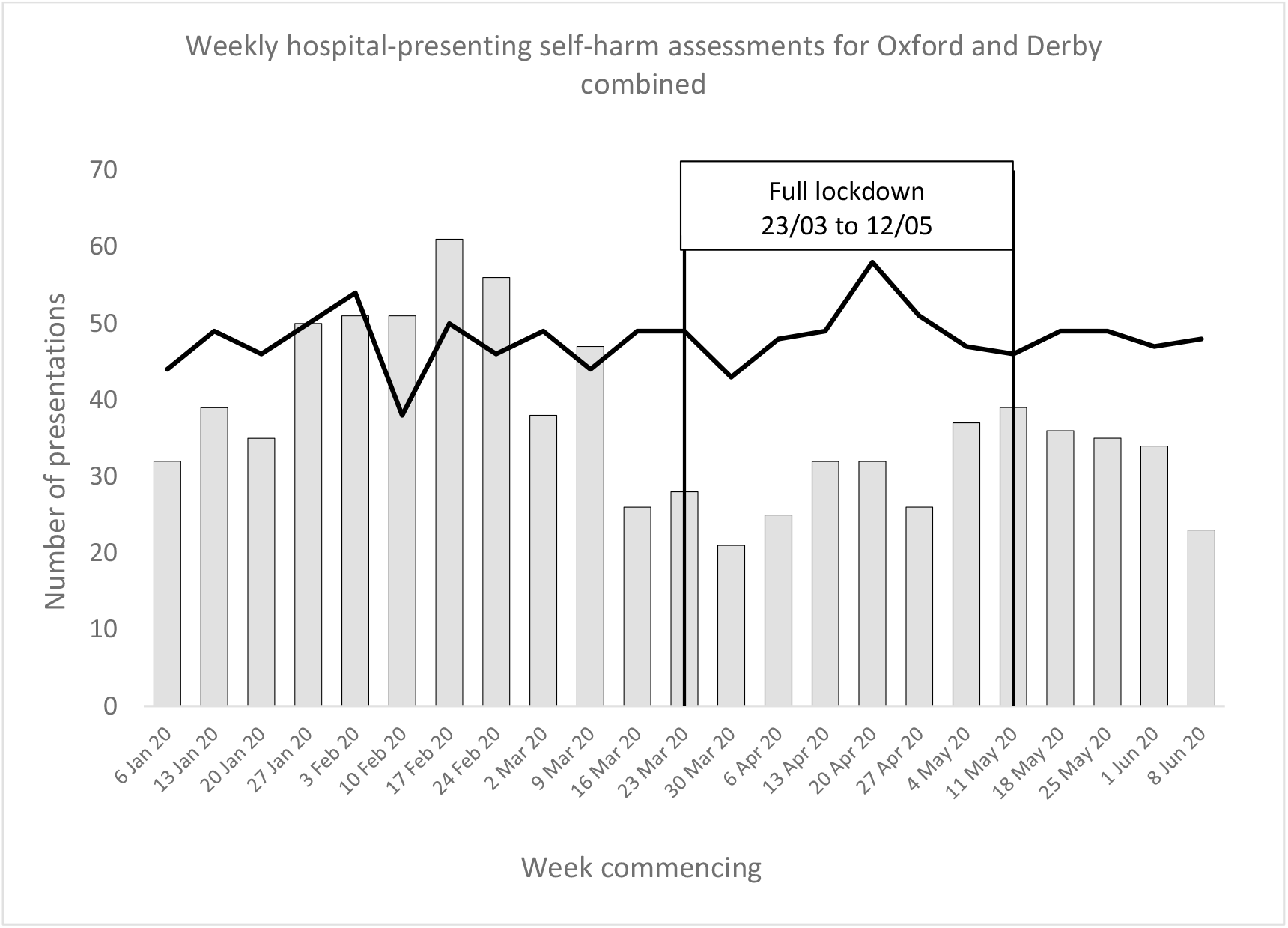
Weekly hospital presentations for self-harm in Oxford and Derby, January 6^th^ – June 14^th^, 2020 and 2019. Data refer to assessed individuals.

During the first 12 weeks following the introduction of lockdown, the average weekly number of self-harm presentations was 30.6% lower than in the pre-lockdown period (6^th^ January to 22^nd^ March 2020), from a mean of 44.2 presentations per week to 30.7 presentations per week, respectively. For the same period in 2019, there was a small, not statistically significant, increase in mean weekly presentations (2.1%). The mean weekly number of episodes during the first 12 weeks following lockdown in 2020 was 37.0% lower than the same period in 2019, from 48.7 presentations per week in 2019 to 30.7 presentations per week in 2020, a mean difference of 18.0 episodes per week (Table 1).

**Table 1.** Mean number of self-harm presentations per week during the period following lockdown relative to the pre-lockdown period and the equivalent periods in 2019. Data refers to assessed presentations.

|  | **Mean number of self-harm assessments per week** |  |  |  |
| --- | --- | --- | --- | --- |
|  | **Pre-lockdown**<br>(January 6<sup>th</sup> - March 22<sup>nd</sup>) | **Post-lockdown**<br>( March 23<sup>rd</sup> - June 14<sup>th</sup>) | **Mean difference (95% CI)** | **P\*** |
| 2020 | **44.2** | **30.7** | 13.5<br>(5.6 to 21.4) | **0.002** |
| 2019 | **47.0** | **48.7** | 1.69<br>(-5.3 to 1.9) | 0.330 |
| **Mean difference (95% CI)** | 2.8<br>(-4.9 to 10.5) | 18.0<br>(13.9 to 22.1) |  |  |
| **P\*** | 0.443 | **<0.001** |  |  |

*\* Using Students t*

A Negative Binomial regression model was used in order to be able to estimate the effect of the 12 weeks of lockdown on presentations compared with before lockdown. The model showed that the number of weekly self-harm presentations following lockdown decreased by 39.6% compared to before lockdown (IRR 0.61, 95% CI 0.46 to 0.78.

### Gender

In terms of gender, the reduction in mean weekly number of presentations after lockdown in 2020 was 11.1 episodes per week (95% CI 5.8 to 16.4) in females compared with a mean reduction of 1.9 episodes per week (95% CI −1.7 to 6.5) in males. The female-to-male presentation ratio changed from 2.0:1 pre-lockdown to 1.5:1 after lockdown. The IRR for males comparing mean weekly numbers to before lockdown was 0.84 (95% CI 0.64 – 1.09) and for females was 0.62 (95% CI 0.51 – 0.76). Thus the incidence in men reduced by an average of 16.2% and in women by 37.8%. Although the reduction was greater in females, the difference was not statistically significantly different (Table 2).

**Table 2.** Incidence rate of presentations to hospital following self-harm in the 12 weeks after the introduction of lock-down compared with pre-lockdown period in 2020: incidence rates (Incidence Rate Ratio and 95% Confidence Interval), by gender, age group and overall method of self-harm.

|  | **IRR** | **95% CI** |
| --- | --- | --- |
| **Gender** |  |  |
| Male | 0.84 | 0.64 – 1.09 |
| Female | 0.62 | 0.51 – 0.76 |
| **Age Group** |  |  |
| 18-34 yrs | 0.56 | 0.45 – 0.70 |
| 35-54 yrs | 0.63 | 0.49 – 0.82 |
| 55+ yrs | 0.72 | 0.43 – 1.22 |
| **Method of self-harm** |  |  |
| Self-poisoning | 0.63 | 0.52 – 0.77 |
| Self-injury | 0.98 | 0.73 – 1.30 |
| Both self-poisoning & self-injury | 0.57 | 0.32 – 1.00 |

### Age Group

The reduction in mean weekly number of presentations after lockdown in 2020 was 11.4 episodes per week (95% CI 6.6 to 16.3) for 18-34 year olds; 4.6 episodes per week (95% CI 1.9 to 7.3) for 35-54 year olds; and 1.6 episodes per week (95% CI - 1.2 to 4.3) for those aged 55 years and over. While the post lockdown reduction in weekly episodes was significant in the 18-34 year and 35-54 year groups, they did not differ significantly from one another.

As seen in Table 2, the reduction in the mean weekly number of self-harm presentations from pre-to post-lockdown in 2020 varied by age group. In 18-34 year old patients, presentations reduced by 43.8%; in the 35-64 year group, they reduced by 36.9% and in those aged 55 years and over the reduction in presentations was 27.9%. In terms of the incidence rate ratios, these differences were not significant between the age groups.

### Self-harm methods

The reduction in mean weekly number of presentations after lockdown in 2020 differed by method of self-harm. For those who self-injured, the mean difference post lockdown was 0.2 episodes per week (95% CI −2.5 to 2.93) and for those who self-injured and overdosed at the same episode, the mean weekly reduction was 1.3 episodes (95% CI −0.1 to 2,80). However, there was a large reduction in the weekly presentations for those who took overdoses: 12.0 episodes per week on average (95% CI 6.4 to 17.6). This differed significantly from those who used any form of self-injury (alone or in combination with an overdose).

There was a difference in the proportion of presentations involving different methods of self-harm before and after lockdown. Presentations involving self-injury alone reduced the least after lockdown (2.4%) whereas self-poisonings reduced by 36.7% and those using both methods reduces by 43.4%, although the numbers in this group were small and the confidence intervals are wide (Table 2).

## DISCUSSION

Considerable concern has been expressed about the potential impacts of the COVID-19 pandemic on mental health (Holmes et al, 2020), especially during the period of lockdown (Brooks et al,2020; Pierce et al, 2020). This study has shown that during the 12-week period following the introduction of lockdown in the UK on March 23^rd^ 2020 there was a very marked reduction in the mean number of weekly presentations for self-harm by individuals aged 18 years and over to two general hospitals involved in the Multicentre Study of Self-harm in England. The reduction was more than 30% compared to the pre-lockdown weeks in 2020 and also the equivalent period in 2019. The reduction appeared to be more marked for presentations involving self-poisoning compared with self-injury. It was greater in 18-34 year olds than in older adults. The reduction in presentations may have begun during the week before lockdown (the forthcoming lockdown having been announced on March 16^th^). It also appeared to persist during the month following the easing of lockdown in England.

We are unaware of any other published data on self-harm presentations to hospitals in England following lockdown, but unpublished data from other hospitals in England also indicate a reduction in presentations following self-harm. An early report from Paris, France, indicated reduced referrals to mental health services for attempted suicide in the city in the first four weeks following lockdown (Pignon et al, 2020). A Spanish study from a general hospital Madrid indicated a major reduction in individuals presenting to the emergency department with suicidal ideation and suicide attempts in March and April following development of the pandemic (Hernandez-Calle et al., 2020). In the Mid-West of the USA a very marked reduction in presentations to emergency departments for suicidal ideation was identified in the month following ‘stay at home’ orders introduced on March 24^th^ 2020 compared with the same period in 2019 (Smalley et al., 2020).

Our findings could have two possible explanations. The first is that self-harm became less common in the community following lockdown. If so, this could have resulted from the impact of the sense of being in it together battling the external threat of the pandemic, in keeping with theoretical explanations for reductions in suicidal behaviour seen during war-time. It could reflect closer living circumstances within families and couples. While in some circumstances this might exacerbate interpersonal problems, in most situations it could be protective, especially for younger people. While lockdown might have increased some stresses, especially in already troubled relationships, it could also have increased cohesion between individuals. Clearly this explanation is unlikely in individuals living alone. Interestingly, a large-scale weekly community survey in the UK in which self-reported data have been collected in a large sample of individuals aged 18 years and over since 23^rd^ March 2020 showed no increase in self-reported self-harm during lockdown (Fancourt et al., 2020), although data for the preceding period are not available for comparison. It is notable that during the period of our study, self-reported depression and anxiety decreased substantially in the survey respondents (Fancourt et al., 2020).

Another potential explanation for our findings is that during the period following introduction of lockdown some individuals who self-harmed avoided going to hospital because of fear of possible exposure to patients with COVID-19 infections. However, the particularly marked reduction in presentations for self-poisoning found in our study period is notable. Most individuals who intentionally self-poison who go to hospital are conveyed by ambulance. When an ambulance is called, the ambulance staff are unlikely to avoid taking patients to hospital, for obvious patient-safety reasons. However, should avoidance of hospital be a contributory factor, in the advent of a further increase in COVID-19 infections consideration should be given to having advertised peripheral clinics that can at least deal with less serious self-harm not requiring the treatment facilities available in general hospitals.

The findings of this study are unlikely to be indicative of what will happen in future, particularly when the likely impacts of the longer-term consequences of the pandemic develop (Gunnell et al., 2020; Reger et al., 2020), especially recession, unemployment and financial problems, all well-known to be associated with increases in suicide and self-harm (Chang et al., 2013; Hawton et al., 2014). However, increases in suicidal behaviour are not inevitable (Gunnell et al., 2020). There is a major need for preventive initiatives to limit the potential impacts of the health crisis leading to increases in suicidal behaviour (Gunnell et al., 2020; Niederkrotenthaler et al., 2020).

For people who present to clinical services having self-harmed there are also issues to address with regard to management. These will include the provision of remote assessments and interventions, and specific therapies directed at helping with the psychosocial consequences of the pandemic.

### Strengths and Limitations

The data for this study were collected through two well-established self-harm monitoring systems (Geulayov et al., 2016), which also allowed us to compare data following lockdown with earlier periods, including in 2019. However, they are based on episodes where the individuals received a psychosocial assessment. In both hospitals the proportion of all self-harm presentations to the emergency departments in which a psychosocial assessment is conducted is high (approximately 75%) especially compared with other hospitals in England (Cooper et al., 2013). We have no reason to believe that this proportion decreased during the period of lockdown, although there was some reduction in the availability of mental health staff in one centre. Also, our findings are similar to those being reported anecdotally from other hospitals in England, in Paris during the first four weeks of lockdown (Pignon et al., 2020), in Madrid during the early stages of the pandemic (Hernandez-Calle et al., 2020) and in presentations of individuals with suicidal ideation to emergency departments in the USA (Smalley et al., 2020).

## CONCLUSIONS

There was a marked reduction in presentations following self-harm in individuals aged 18 years and over to two major general hospitals in England during the first three months following lockdown due to the COVID-19 pandemic. These findings, which are consistent with findings from other countries, might have multiple explanations. They cannot be taken as indicative of what will happen with regard to future levels of self-harm, especially as the longer-term impacts of the pandemic play out. This is a major reason for maintaining high quality surveillance of self-harm presentations to hospitals.

## Data Availability

The data from this stuy are not available as our approval from Health Research Authority Confidentiality Advisory Group under Section 251 of the NHS Act 2006 (which allows collection of patient data without patient consent)does not allow data sharing

## Contributors

KH and DC were responsible for study conception and design, and interpretation of the results. DC and GG were responsible for data analysis. LB, FB, JN and KW acquired the data. KH drafted the report, which all authors critically revised for intellectual content. All authors approved the final report and are accountable for all aspects of this work. KH supervised the study and is the guarantor.

## Acknowledgements

The study was funded by the Department of Health and Social Care. KH is supported by Oxford Health NHS Foundation Trust.

We thank the clinicians and the research staff in both centres for their support with the data collection. The authors from Derby would like to thank Jessica Pearson and Phyllis Leung (Research Assistants).

Role of the funding source

The Department of Health and Social Care had no role in study design, data collection, analysis, and interpretation of data, or in the writing of the report, and in the decision to submit the paper for publication.

